# Demographics, Epidemiology, Mortality, and Difficult-To-Treat Resistance Patterns of Bacterial Bloodstream Infections in the Global United States Military Health System from 2010-2019: A Retrospective Cohort Study

**DOI:** 10.1101/2024.10.02.24314780

**Authors:** Alexander C Vostal, Melissa Grance, John H Powers, Sameer S Kadri, Sarah Warner, Uzo Chukwuma, Carlos Morales, Charlotte Lanteri, M Leigh Carson, Beth Poitras, Nicholas Seliga, Dean Follmann, Jing Wang, Edward Parmelee, Katrin Mende

## Abstract

**Objective:** To describe demographics, causative pathogens, hospitalization, mortality, and antimicrobial resistance of bacterial bloodstream infections (BSIs) among beneficiaries in the global U.S. Military Health System (MHS), a single-provider healthcare system with 10-year longitudinal follow-up.

**Design:** Retrospective cohort study

**Setting:** Clinical and demographic data collected from the MHS Data Repository and collated with microbiological data obtained from the Defense Centers for Public Health-Portsmouth.

**Participants:** 12,748 MHS beneficiaries diagnosed with 15,357 bacterial BSIs (2010-2019).

**Main Outcome(s) and Measure(s):** Demographic data and diagnosis codes preceding BSI episodes and during hospitalizations were collected. Inpatient admission data identified acute clinical diagnoses, intensive care unit (ICU) admission, and mortality. BSI pathogens were evaluated for antimicrobial resistance, including difficult-to-treat resistance (DTR). Crude mortality trends were assessed.

**Results:** The decade analyzed included 15,357 BSI episodes in 12,748 patients; 6,216 patients (48.8%) were ≥65 years and 83.7% of episodes had ≥1 comorbidity (12,856 of 15,357). Approximately 29% of episodes with hospitalization required ICU admission and ∼34% had concurrent urinary tract infections. Pathogen distribution was 53% and 47% for Gram-positive bacteria and Gram-negative bacilli (GNB), respectively. Inpatient mortality was 4.4%, and at one year was 23.4%; 0.5% (16 of 2,977) of deaths were associated with DTR GNB. Among an average 8,145,778 individuals receiving care annually in the MHS, annual rates of overall BSI, methicillin-resistant *Staphylococcus aureus*, vancomycin-resistant *Enterococcus* spp., and DTR GNB BSI were 18.9, 1.30, 0.25, and 0.05 per 100,000 beneficiaries, respectively. Over the decade, annual mortality did not significantly increase for any pathogen and decreased by ∼3% for lactose-fermenting GNB BSI (p=0.048).

**Conclusions:** In the global U.S. MHS, mortality burden associated with BSI was substantial (approximately 1 in 4 dying at 1 year), relatively unchanged over a decade, and associated with older age and comorbidities. First-line treatment options remained available for 99.7% of BSIs. Population-level improvements in BSI survival might be maximally influenced by focusing on prevention, early detection, prompt antibiotics, and other novel therapies not contingent on *in vitro* activity.

**Summary Box:** **What is already known on this topic:**

- Bloodstream infections (BSIs) are associated with high healthcare burden and poor patient outcomes, including high mortality.
- Modeling data based on assumptions suggest that mortality associated with antimicrobial-resistant pathogens is increasing.

**What this study adds:**

- Among Military Health System (MHS) beneficiaries, overall and difficult-to-treat antimicrobial-resistant BSIs averaged an annual rate of 18.9 and 0.05 per 100,000 beneficiaries, respectively.
- Over a decade, mortality did not increase annually for any BSI group, while lactose-fermenting Gram-negative BSI mortality decreased (∼3%) and 50% of BSIs associated with deaths at 1-year occurred >42 days after BSI diagnosis.
- Bacterial BSI deaths in MHS are often associated with advanced age (74% ≥65 years) and comorbidities (97% with ≥1 comorbidity), rather than absence of first-line antimicrobial treatment options.

## INTRODUCTION

Bloodstream infections (BSIs) are associated with high healthcare burden and poor patient outcomes. Inpatient mortality rates from healthcare- or community-associated BSIs range from 18% to 48%.^1–5^ Worldwide, the most common pathogens associated with BSIs in adults are *Staphylococcus aureus* and Enterobacterales.^6,7^ Longitudinal impact on BSI patient outcomes with antimicrobial-resistant (AMR) and susceptible pathogens needs further evaluation.

Modeling data from a decade ago suggest AMR pathogens will kill more patients than cancer by 2050.^8^ Models are based on assumptions, not real-world evidence and AMR incidence varies per definitions used. Studies evaluating AMR often assess resistance to individual or various combinations of drugs/classes. Kadri et. al.^9^ posed a more clinically-relevant definition termed “Difficult-to-Treat” resistance (DTR) in 2018 that better correlates with mortality by evaluating how many drugs remain biologically active *in vitro*. DTR for Gram-negative bacilli (GNB) is defined as *in vitro* resistance to all three classes of first-line antibiotics, leaving drugs with questionable efficacy and greater toxicities. In U.S. hospitals, <1% of GNB BSI between 2009-2013 were DTR, with *Pseudomonas aeruginosa* and *Acinetobacter baumannii* as main contributors (2.3% and 18.3% of DTRs).^9^ Independent of AMR, in-hospital mortality in patients with severe sepsis decreased from 45.0% to 27.8% (1993-2008), while organism-specific mortality risks remained relatively unchanged,^10^ suggesting patient factors other than *in vitro* activity impact BSI mortality and warrant further evaluation.

We describe the epidemiology of bacterial BSI within the global U.S. Military Healthcare System (MHS), where single-provider universal healthcare and longitudinal follow-up are provided for active-duty and retired U.S. service members and beneficiaries. This 10-year assessment of BSIs evaluated characteristics, incidence of infections and etiologies, resistance patterns, and longitudinal mortality with BSIs.

## METHODS

### Study design

This was an observational retrospective cohort study of all recorded individuals receiving healthcare within the MHS who developed bacterial BSIs (1/1/2010–12/31/2019). The MHS is a global single-provider system delivering healthcare to 9.6 million active-duty, reserve-component and retired U.S. military personnel and their dependents at 45 hospitals (31 in the USA) and 566 ambulatory care and occupational health centers (466 in the USA).^11,12^ Microbiological data obtained from the Defense Centers for Public Health-Portsmouth were collated with clinical and demographic data from the MHS Data Repository. Patient identifiers from blood cultures were aggregated with clinical data from the MHS Data Repository, which prospectively captures clinical and microbiological data from individuals receiving care at military treatment facilities (MTF) or contracted civilian healthcare facilities. This study (IDCRP-108) was approved by the Uniformed Services University (USU) Institutional Review Board (IRB). We followed STROBE guidelines for reporting observational studies.^13^

### Study Population and Definitions

The study population consisted of adults (≥18 years) with clinical encounters (**Supplemental Material**) or recorded care episodes in clinical settings within the MHS or contracted civilian healthcare facilities. This includes U.S. service members (active-duty and retired) and their beneficiaries, and civilians receiving emergency care from MTFs. Due to a lack of longitudinal clinical data, we excluded civilian individuals without MHS medical benefits from final descriptive analyses. BSI episodes were defined by single positive blood cultures, with select, clinically-relevant bacterial pathogens (**Supplemental Table 1**), with associated clinical encounters. Episodes without associated clinical encounters or missing demographic data were excluded (**Figure 1a**).

**Figure 1.**
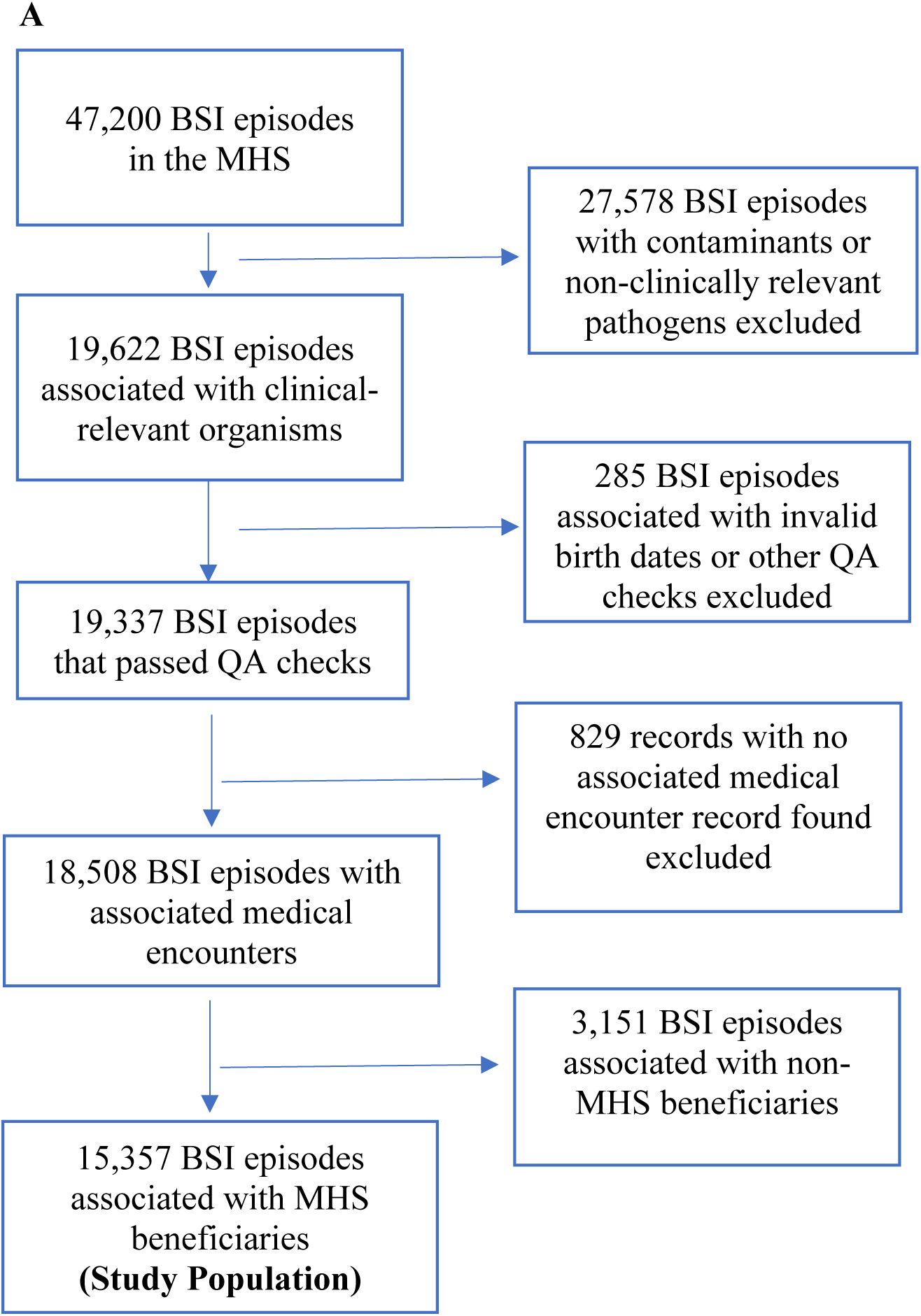
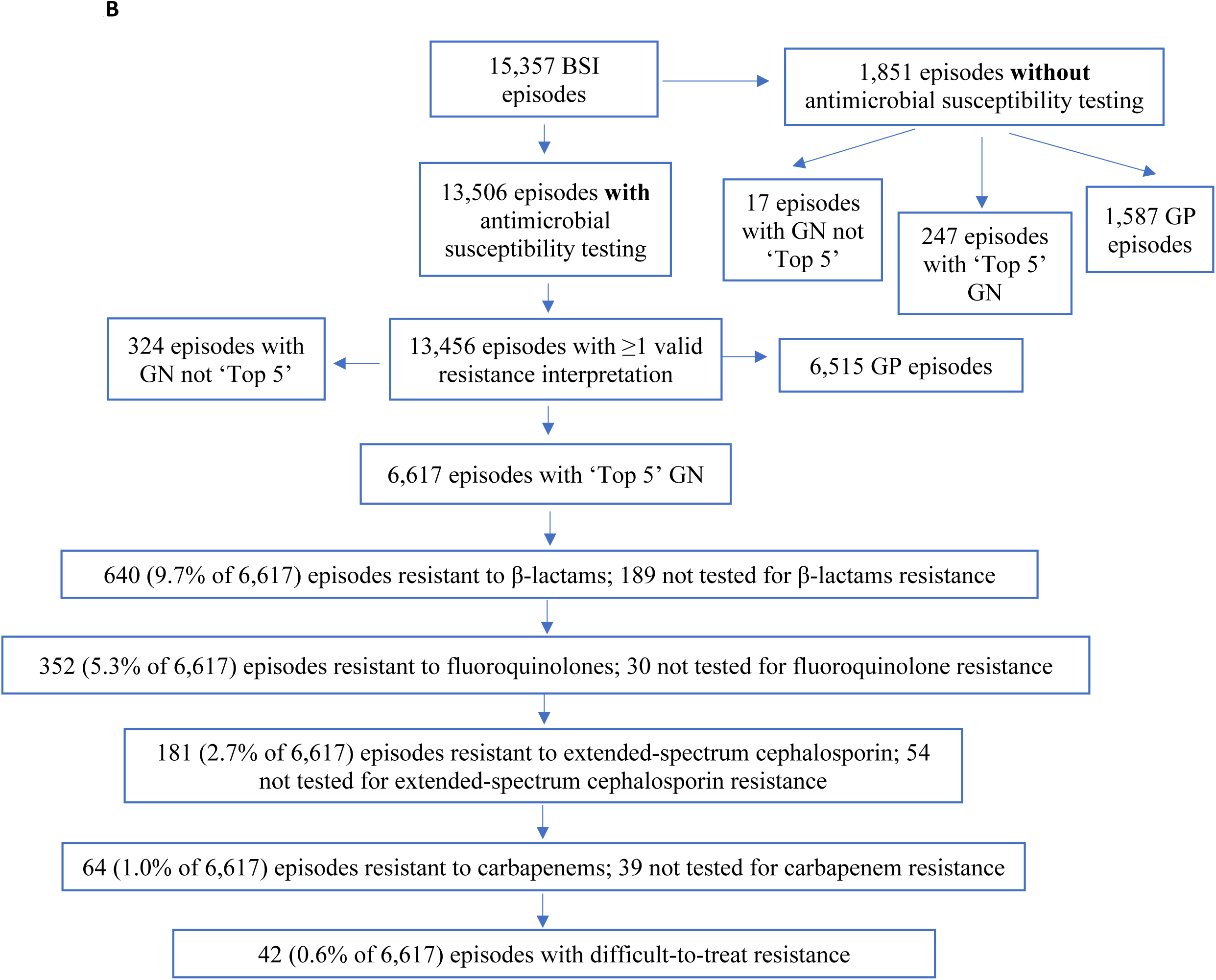
Flow diagram for: (**A**) Bloodstream infection (BSI) episodes diagnosed between 2010-2019 included in the analysis. (**B**) Pathogens and antimicrobial susceptibility testing. ‘Top 5’ represents the most numerous and clinically relevant pathogens (i.e., *E. coli*, *Acinetobacter* spp., *K. pneumoniae*, *P. aeruginosa*, and *Citrobacter* spp.). Pathogens not in the ‘Top 5’ include *Stenotrophomonas* spp. and *Enterobacter* spp. GN pathogens assessed for DTR were those that had susceptibility testing for three classes of first-line antibiotics (i.e., carbapenems, extended-spectrum beta-lactams, or fluoroquinolones). Antibiotic resistance information for GP organisms is in **Supplemental Figure 2**. DTR – difficult to treat resistance; GN – Gram negative; GP – Gram-positive; MHS – Military Health System; QA – quality assurance

Demographic, clinical, and microbiological data were collated for patients having BSIs associated with clinically-relevant organisms (**Supplemental Table 1; Supplemental Figure 1**). Comorbidities were determined via International Classification of Diseases (ICD) 9/10 codes associated with patients’ electronic medical records (EMRs), ≥7 days preceding positive blood cultures. ICD9/ICD10 codes were subcategorized and used to calculate updated Charlson Comorbidity Indices (uCCI).^14,15^ Comorbidities were grouped using an amalgamation of Clinical Classifications Software diagnosis mapping categories.^16^ Clinical data, length of stay, and inpatient mortality data were abstracted from inpatient admission records with dates corresponding to BSIs, including episodes preceding admission by <u>≥</u>7 days. ICD9/10 Trauma Procedure codes ≤30 days prior to BSIs were identified. For individuals with multiple BSIs, comorbidity data were collated for each unique BSI to ensure mutual exclusivity.

### Microbiology

Bacterial identification and antimicrobial susceptibility testing were performed per institutional guidelines and methods at MTF laboratories (**Supplemental Material**).

Clinical and Laboratory Standards Institute minimum inhibitory concentrations (MIC) interpretive criteria were utilized to determine susceptibility profiles.^17^ Those with “intermediate” *in vitro* results were classified as “resistant” for this analysis. Organisms were considered resistant to classes of antimicrobials if they were resistant to all drugs tested in that class (antimicrobials listed may not include each member of the class tested at each laboratory).

*In vitro* susceptibilities for GNB used the DTR algorithm^9^ defined as antimicrobial resistance to all three classes used for safe and effective treatment of serious GNB infections (carbapenems, fluoroquinolones, and extended-spectrum cephalosporins; see **Supplemental Material**). Antibiotic resistance for Gram-positive bacteria (GPB) focused on patterns of clinical relevance (e.g., methicillin resistance in *S. aureus* and vancomycin resistance in *Enterococcus* spp.). For GPB, we defined DTR as isolates simultaneously resistant to vancomycin, linezolid, and daptomycin given recommendations for their use in resistant GPB infections.^18,19^

For final analyses, microbiological results were restricted to the 15 most frequently recovered bacterial species, then further subcategorized into four groups reflecting shared clinical pathogenesis: 1) Lactose-fermenting GNB (including *Proteus* spp.), 2) *Streptococcus* and *Enterococcus* species, 3) *S. aureus*, and 4) Non-lactose-fermenting GNB (**Supplemental Table 1**).

### Statistical Analysis

Descriptive statistics are presented for demographic and microbiological data. Incidence was calculated using yearly BSI (only clinically-relevant organisms) count per total annual beneficiaries. Annual trends for incidence and mortality per BSI episode based on bacterial groups were evaluated (see **Supplemental Material** for statistical methods).

### Patient and Public Involvement

This is a registry-based study, which was granted a waiver of informed consent by the USU IRB. The patients in this setting are active-duty military personnel and their relatives. The corresponding author (LCDR Alexander Vostal) and one of the included investigators (LTC Charlotte Lanteri) are active-duty service members whose relatives are also beneficiaries of the Military Health System. Both provided input into the design, conduct and analysis of the study.

## RESULTS

### Study population

#### Total BSIs

Between 2010 and 2019, an average of 8,145,778 individuals per year received care within the MHS, and 15,261 individuals had 18,508 BSI episodes (**Figure 1a**). Among those, 12,748 (83.5%) were MHS beneficiaries (or eligible for MHS services) with 15,357 BSI episodes (average annual rate of 18.9 per 100,000 beneficiaries) included in the analyses. The majority were male (54.8%), >65 years (48.8%), and retired uniform service members (42.9%) (**Table 1**). BSIs were primarily diagnosed at facilities within the continental USA (**Supplemental Table 2**).

**Table 1.** Demographics of Military Health System (MHS) beneficiaries who Developed a Bloodstream Infection.

| **Demographics** | **Patients**<br>**No. (%)**<br>**(N=12,748)** |
| --- | --- |
| *Age Group (years)<sup>a</sup>* |  |
| 18-25 | 1,162 (9.1) |
| 26-35 | 990 (7.8) |
| 36-45 | 821 (6.4) |
| 46-64 | 3,559 (27.9) |
| ≥65 | 6,216 (48.8) |
| *Sex* |  |
| Female | 5,767 (45.2) |
| Male | 6,981 (54.8) |
| *Race* |  |
| African American | 2,050 (16.1) |
| Asian or Pacific Islander | 929 (7.3) |
| Caucasian | 7,325 (57.5) |
| Other | 2,444 (19.2) |
| *Type of MHS Beneficiary<sup>a</sup>* |  |
| Active duty | 1,587 (12.4) |
| Dependents | 5,368 (42.1) |
| Military retirees | 5,465 (42.9) |
| Other | 328 (2.6) |

<sup>a</sup> For individuals with multiple BSI episodes, age and MHS beneficiary type data collected from the first episodes are presented.

The pre-BSI median uCCI for MHS beneficiaries was 3.0, indicating moderate comorbidities (**Supplemental Table 3**). Comorbidities were common with the most frequent being chronic pulmonary disease (49.6%), diabetes (45.2%), peripheral vascular disease (34.9%), malignancy (34.1%, excluding non-melanoma malignant skin neoplasms), and renal disease (34.3%; **Supplemental Table 3**). Sixteen percent did not have comorbidities. Trauma diagnoses preceded 11.6% (1,786 of 15,357) of episodes (**Supplemental Results**). Thirty-nine individuals had BSIs with DTR GNB (3 with 2 DTR episodes) and were older (54% were ≥46 years) with a median uCCI of 5. No patients had DTR GPB infections. Demographics on the level of the BSI episode are in **Supplemental Table 4**.

### Bacterial Pathogens

#### Microbial Etiologies

*Escherichia coli* (29.9%; 4,585 of 15,357) and *S. aureus* (19.8%; N=3,040) were the most common pathogens, yet *Enterococcus* and *Streptococcus* species combined (including β-hemolytic *Streptococcus* and *Streptococcus* viridans group) accounted for 33% (N=5,072; **Table 2**; **Figure 2A**). *P. aeruginosa* and *Acinetobacter* spp. represented 3.8% (N=580) and 1.2% (N=186), respectively.

**Figure 2.**
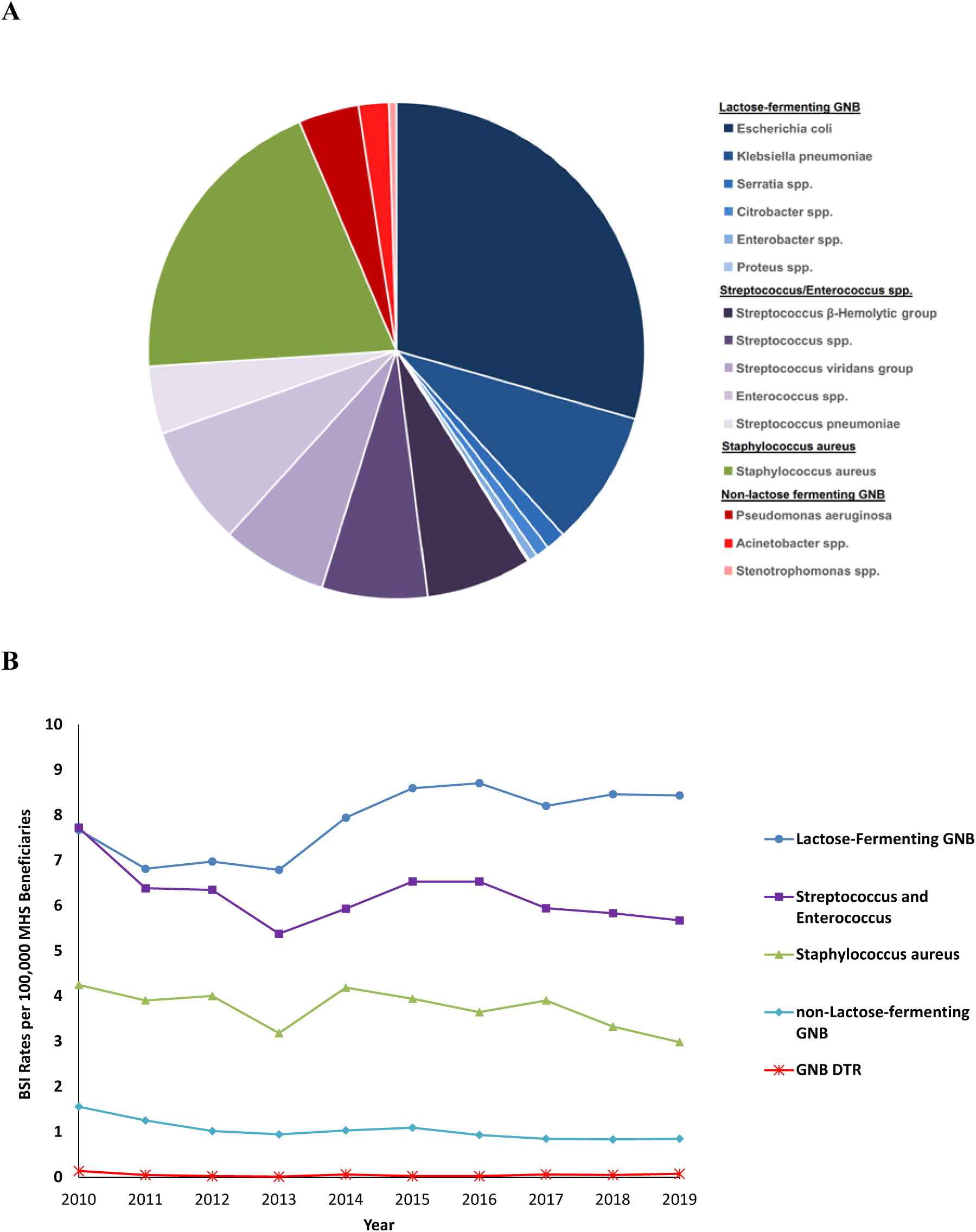
Bacteria associated with bloodstream infections (BSI). A) Distribution of total bacteria; B) BSI rate per 100,000 Military Health System (MHS) beneficiaries per year by classification group. There were no DTR Gram-positive bacteria. DTR – difficult-to-treat resistance; GNB – Gram-negative bacilli

**Table 2.** Frequency of Bacterial Species associated with Bloodstream Infection (BSI)

| **Bacterial Species** | **Number of all BSI Episodes** | **Total Number of Patients with BSI Episodes<sup>a</sup>** |
| --- | --- | --- |
| **Total** | **15,357** | **12,748** |
| *Escherichia coli* | 4,585 | 4,355 |
| *Staphylococcus aureus* | 3,040 | 2,825 |
| *Klebsiella pneumoniae* | 1,369 | 1,272 |
| *Enterococcus* spp. | 1,258 | 1,154 |
| *Streptococcus* β-hemolytic group | 1,085 | 1,018 |
| *Streptococcus* spp. | 1,035 | 1,015 |
| *Streptococcus viridans* group | 1,006 | 989 |
| *Streptococcus pneumoniae* | 688 | 672 |
| *Pseudomonas aeruginosa* | 580 | 556 |
| *Serratia* spp. | 194 | 192 |
| *Acinetobacter* spp. | 186 | 180 |
| *Citrobacter* spp. | 144 | 141 |
| *Enterobacter* spp. | 90 | 87 |
| *Stenotrophomonas* spp. | 78 | 75 |
| *Proteus* spp. | 19 | 19 |

<sup>a</sup> Patient counts are not mutually exclusive

Lactose-fermenting GNB BSIs were the most frequently identified grouping (41.7%; 6,401 of 15,357). Non-lactose-fermenting GNB were the lowest proportion (5.5%; N=844; **Figure 2A**). The highest average annual incidence over the decade was lactose-fermenting GNB at 7.8 cases per 100,000 MHS beneficiaries, followed by *Streptococcus*/*Enterococcus* spp. (6.2 per 100,000) and *S. aureus* (3.7 per 100,000) (see **Figure 2B** for annual rates). The lowest average annual rate was non-lactose-fermenting GNB, including *Acinetobacter* (1 per 100,000), 7.8-fold lower than lactose-fermenting GNB.

#### *In Vitro* Antimicrobial Resistance

Among 15,357 BSIs, 13,456 (88%) had available susceptibility results with 49% (6,617 of 13,456) diagnosed with one of five most common, clinically-relevant GNB (*E. coli*, *K. pneumoniae*, *P. aeruginosa, Acinetobacter* spp. and *Citrobacter* spp.,), 2% (N=324) diagnosed with another GNB and 48% (N=6,515) diagnosed with GPB (**Figure 1b; Supplemental Figure 2**).

Within the top five GNBs BSIs over the decade, 640 (9.7% of 6,617) were resistant to beta-lactams (**Figure 1b**). In a decision stepwise approach, 5.3% were resistant to fluoroquinolones, 2.7% were resistant to extended-spectrum cephalosporins, 1.0% were resistant to carbapenems, and 0.6% (42 of 6,617; 0.3% of 15,357 total BSIs) were DTR (average BSI annual rate: 0.05 per 100,000 MHS beneficiaries; see **Figure 2B** for annual rates). Among 42 BSIs with DTR pathogens, 19 (45.2% of DTR/10% of total *Acinetobacter*/0.1% of all organisms isolated) isolated *Acinetobacter* spp., 14 (33.3%/2.4%/0.09%) *P. aeruginosa*, 8 (19.0%/ 0.6%/0.05%) *K. pneumoniae*, and 1 (2.4%/0.02%/0.006%) *E. coli*. **Supplemental Table 5** provides resistance data for GNB BSIs (N=3,707) with susceptibility testing against first-line DTR antimicrobial class assessments.

GPBs included methicillin resistance in 36.0% of *S. aureus* (1,057 of 2,936; average annual rate: 1.30 per 100,000 beneficiaries) and vancomycin resistance in 17.3% of *Enterococcus* spp. (205 of 1,187; average annual rate: 0.25 per 100,000 beneficiaries). (**Supplemental Table 5**). *S. aureus* vancomycin and daptomycin resistance among were low (N=4, 0.1%; N=2, 0.5%, respectively). No GPB met DTR criteria. *Streptococcus* spp. (excluding β-hemolytic *Streptococcus* spp.) with extended-spectrum cephalosporin resistance ranged between 2.1-5.7% (**Supplemental Results)**.

### Patient outcomes

Nearly all BSIs (98.8%; 15,180 of 15,357) had associated hospitalizations with 21,497 unique hospital admissions (includes emergency room admissions) for a median hospital duration of 5 days (range: 1-1,020 days). Excluding emergency room admission, 28.9% of 15,588 hospitalizations required transfer to intensive care units (ICUs). Inpatient diagnoses of septicemia (except in labor), urinary tract infection (UTI), pneumonia, and skin and subcutaneous tissue infection were observed in 48.5%, 33.6%, 17.9%, and 11.3% of 21,497 unique hospital admissions, respectively. Endocarditis was diagnosed with 3.5% of hospital admissions. Acute (including unspecified) renal failure, respiratory failure, and shock were diagnosed in 25.9%, 12.4%, and 9.5% of hospital admissions, respectively. Central venous catheter infections accompanied 3.2% of BSI-associated hospitalizations.

In a per patient analysis, inpatient mortality was 4.4% (937 hospital deaths in 21,497 admissions; median 11 days [range: 1-652 days] to death post-admission) and 23.4% (2,977 of 12,748; median 43 days [range: 1-362 days] from BSI episode) of individuals died within 1 year of a BSI episode with 5% having polymicrobial infections. Sixteen (0.5%) individuals who died had DTR BSIs (2 with polymicrobial infections). Pneumonia/respiratory failure was the most frequent diagnosis associated with mortality. UTIs contributed <1%. Individuals who died were predominantly older (74% ≥65 years) and 97% had ≥1 comorbidity (**Supplemental Table 6**).

Analyzing per BSI episode, a total of 24.3% (3,725 of 15,357) were associated with mortality within 1 year of BSI (23 [0.6% of 3,725] with DTR). There was a significant downward trend in 1-year mortality with all BSIs from 27% in 2010 to 18% in 2019 (p=0.024). *Streptococcus/Enterococcus* spp. BSIs (35.1%; 1,306 of 3,725) were most frequently associated with death, then lactose-fermenting GNB (33.2%; N=1,236), *S. aureus* (24.0%; N=895) and non-lactose-fermenting GNB BSIs (7.7%; N=288). Nine lactose-fermenting GNB and 33 non-lactose-fermenting GNB BSI episodes met DTR criteria with 0.6% (7 of 1,236) and 5.6% (16 of 288) of episodes associated with death, respectively (**Figure 3**). Trend analysis for bacterial groups demonstrated a significant decrease in mortality for lactose-fermenting GNB from 22% in 2010 to 14% in 2019 (p=0.048). Mortality did not significantly increase for any bacterial BSI group.

**Figure 3.**
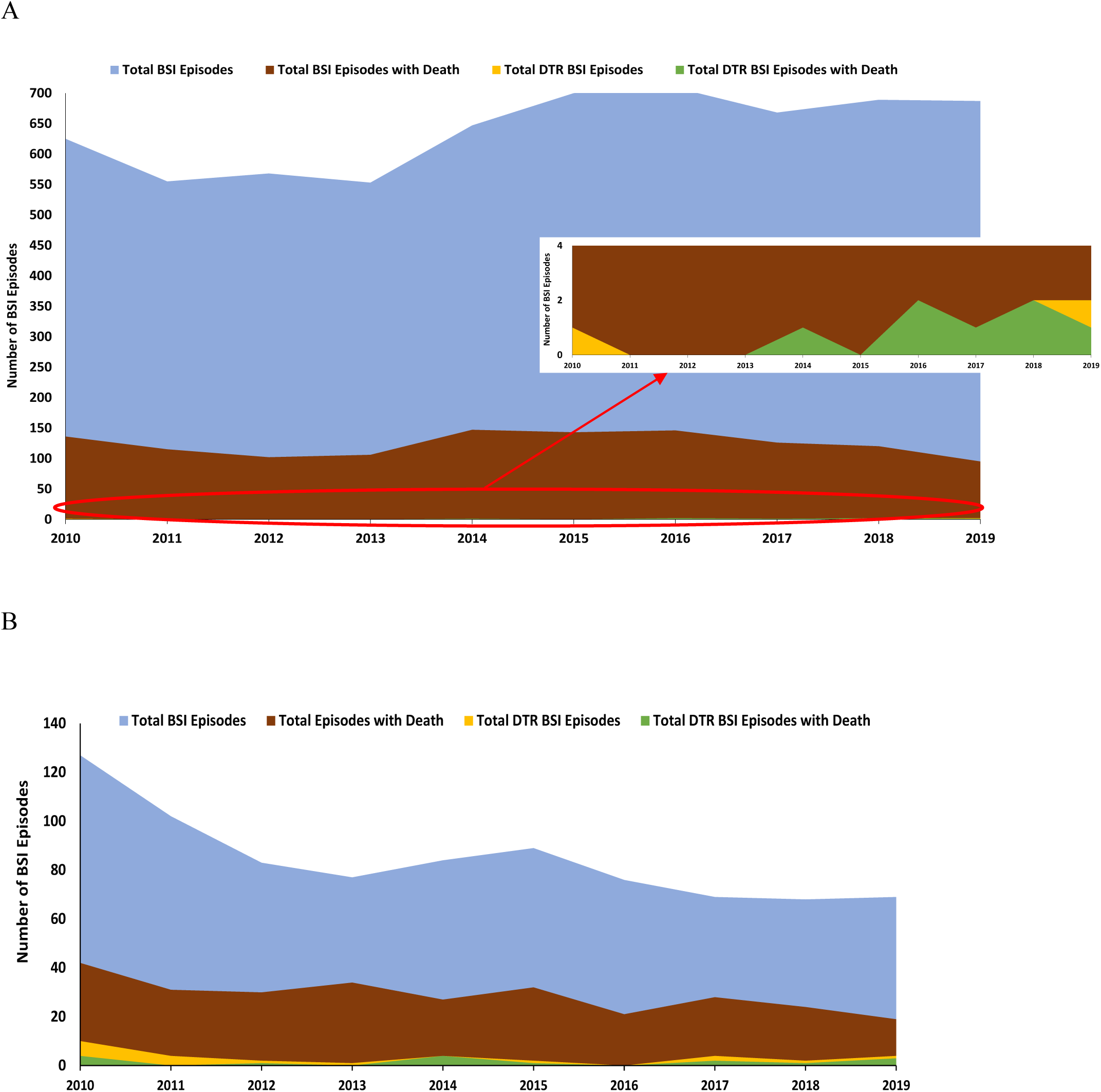
Total number of (A) lactose-fermenting Gram-negative bacilli and (B) non-lactose-fermenting Gram-negative bacilli bloodstream infection (BSI) episodes and deaths, including occurrence of difficult-to-treat resistance (DTR). Of the 16 DTR BSI episodes with deaths, one patient had two different DTR GNB (*Pseudomonas aeruginosa* and *Klebsiella pneumoniae*) on the same day and one patient had a polymicrobial BSI (DTR *P. aeruginosa* plus three susceptible pathogens: *Enterococcus* spp., *K. pneumoniae*, and *Staphylococcus aureus*)

## DISCUSSION

We observed 15,357 BSI episodes in 12,748 MHS beneficiaries over 10 years in a global single-provider healthcare system with longitudinal follow-up and uniform data capture, including survival, and classified resistance as clinically-relevant DTR. Among hospitalized patients, 28.9% had ICU admissions and 4.4% died during the admission, demonstrating BSI’s substantial burden. Despite most military personnel being young and healthy, males aged >46 years old and individuals with COPD largely comprised this cohort, suggesting BSIs are not randomly distributed. UTIs were diagnosed twice as frequent as pneumonia during hospital admission.

*E. coli* and *S. aureus* BSIs were most common. DTR pathogens were observed in 3 per 1000 or 39 total patients, remaining low over time, with 99.7% of infections susceptible to currently available antimicrobials. Among patients with BSIs who died, 0.5% had an BSI associated with DTR pathogens Resistance to vancomycin and daptomycin in GPB, specifically methicillin-resistant *S. aureus* or vancomycin-resistant *Enterococcus* spp., remained low. No patient had DTR GPBs. Over the decade, there was a significant downward trend in mortality for overall BSIs and those with lactose-fermenting GNB. There was no significant increase in BSI mortality for any bacterial group. Approximately half of DTRs were *Acinetobacter*, yet these were uncommon, occurring in 1 per 1000 or 19 patients. DTRs were rare, occurring in 6 per 100,000 or 1 patient with *E. coli* BSIs and <1% of BSI episodes associated with mortality among DTRs. We utilized a conservative approach by classifying pathogens with intermediate resistance as resistant.

We observed low incidence (<5.0%) of carbapenem resistance in GNBs. A substantial portion of multidrug-resistant organisms were non-lactose fermenters, such as *Acinetobacter* spp. and *P. aeruginosa*. As hospitalizations with non-lactose-fermenting BSIs had the lowest associated mortality, this suggests mortality may be impacted by factors other than *in vitro* resistance.

We studied BSIs as this allowed accurate diagnosis of both disease and microbial etiologies. Although our study was not structured to confirm BSI sources, we did evaluate associated recorded infections. A recent review suggested UTI as the most common source of *E. coli* BSIs.^20^ consistent with our finding of approximately one-third of individuals diagnosed with UTI during inpatient BSI admissions. However, <1% of BSI episodes with mortality had associated UTI diagnoses. As UTIs are more common in females,^21^ higher rates of lactose-fermenting GNB BSIs among females in our study may reflect higher UTI rates.

The evidence from our study points to substantial mortality in patients with susceptible infections, especially associated with respiratory failure/pneumonia. The burden of susceptible infections is often less discussed when focusing on *in vitro* AMR instead of patient outcomes. Poor outcomes may be due to lack of point of care diagnostics, and disordered immune responses independent of antimicrobial susceptibility. A decade after predictions of more deaths with AMR than cancer by 2050,^8^ our data are consistent with current real-world European data showing regularly lower AMR rates than modeling predictions. Recent data also demonstrate decreasing trends of AMR phenotypes at U.S. Veterans Affairs medical centers.^22–28^ *Acinetobacter* outbreaks were previously reported in MTFs (102 patients over 2 years);^29^ however, these personnel were largely injured in Iraq during military conflict, demonstrating resistance can be episodic and does not necessarily continuously increase over time. Our study identified a total of 180 patients with *Acinetobacter* infections over the succeeding decade. The low occurrence of DTRs may reflect effective stewardship practices.

The strengths of this study are that the global MHS includes all patient types, including older patients with co-morbidities, and that the universal care utilized within the MHS reduces the potential for outcomes differences that arise when there are disparities in healthcare systems. Studies evaluating the burden of infections including AMR often are predominantly single-center, cross-sectional studies, use heterogenous definitions of resistance, and/or lack long-term outcomes data. As AMR has been characterized as a national security issue, it is important to evaluate the impact in the MHS specifically.^30^ Although most data were from U.S. sites, military personnel deploy globally and may be admitted to U.S. MTFs post-deployment. Data collection in single-provider systems is more uniform and complete, allowing longitudinal assessments, including mortality. Prior studies examining BSI mortality generally restrict to either inpatient or ≤30-day mortality,^31–37^ and while there are studies examining long-term mortality, GNB resistance is not assessed.^38,39^ We examined 1-year mortality as prior studies show patients continue to have increased long term mortality post-infection.^40^ We also evaluated all relevant records, and unlike other BSI-focused studies, we utilized patient characteristics and assessed outcomes. The microbiological data was that seen by clinicians in clinical practice.

Our study includes limitations inherent with retrospective use of large, non-centralized clinical datasets for analyses (e.g., not all isolates collected were tested for resistance to every drug). Mortality was our sole outcome as other outcome data (e.g., residual disability) were not available and further analyses on attributable cause of death are needed. Although the MHS provides “all encompassing” care for beneficiaries, the transient nature of military service limits beneficiary status and accompanying clinical data for individual patients, but still provides accurate data for the care received within MHS as a whole.

These results also have implications for research. Epidemiological studies should focus on patient outcomes including disease from susceptible organisms rather than focusing solely on AMR. As BSIs were caused by relatively few types of bacteria in specific populations, studies should focus on targeted prevention strategies in specific patient types at greatest risk (e.g., older persons with comorbidities). Since patient factors independent of *in vitro* biologic activity of drugs may be important, future research should include prevention (vaccines and non-pharmacological interventions) and host-directed therapies, including microbiome and immunomodulator therapies.

This study examined the microbiological epidemiology and clinical features of bacterial BSIs over a decade within a comprehensive global healthcare system. Our next step is to characterize both military and civilian specific risk factors for development and survival of BSI, including assessment of attributable mortality. This study shows that further research should focus on patient outcomes, including morbidity and mortality, demonstrating greater impact in patients infected with susceptible pathogens and the need for better therapies patients who are most at risk of poor outcomes independent of causative pathogen resistance. Such patients are often excluded from clinical trials testing new drugs.^41^ Setting of microbiological breakpoints should also be based on patient outcomes as “resistance” to providers implies worse patient outcomes. The goal should be improving patient outcomes for all patients and resources should be in line with this objective, rather than focusing only on AMR.^41^ Further analyses into risk factors and attributable outcomes are warranted and may lead to the development of effective therapeutic and prevention strategies for BSI in patients most at risk of poor outcome regardless of pathogen.

## Data Availability

Data for this study are available from the Infectious Disease Clinical Research Program (IDCRP), headquartered at the Uniformed Services University of the Health Sciences (USU), Department of Preventive Medicine and Biostatistics. Review by the USU Institutional Review Board is required for use of the data collected under this protocol. Furthermore, the data set includes Military Health System data collected under a Data Use Agreement that requires accounting for uses of the data. Data requests may be sent to: Address: 6270B Rockledge Drive, Suite 340, Bethesda, MD 20817.

## Acknowledgments

We would like to thank Martin G. Ottolini, MD, for his contributions to this study.

## Contributors

All authors contributed to the study concept and design, and the acquisition, analysis, or interpretation of data. ACV, JHP, SSK, UC, CM, CL, DF, EP, and KM contributed to study design. Data were obtained by MG, UC, BP, NS, and EP and analysis was conducted by MG and JW. ACV, JHP, SSK SW, MLC, NS, and KM contributed to data interpretation. ACV wrote the first manuscript and MLC (technical writer), JHP, and KM revised the drafts. All authors critically reviewed and approved the final version. The corresponding author (ACV) attests that all listed authors meet authorship criteria and that no others meeting the criteria have been omitted. ACV had full access to all the data in the study and takes responsibility for the integrity of the data and the accuracy of the data analysis.

## Transparency

The lead author (ACV, manuscript guarantor) affirms that the manuscript is an honest, accurate, and transparent account of the study being reported; that no important aspects of the study have been omitted; and that any discrepancies from the study as planned (and, if relevant, registered) have been explained.

## Data Sharing Statement

The Corresponding Author has the right to grant on behalf of all authors and does grant on behalf of all authors, a worldwide license to the Publishers and its licensees in perpetuity, in all forms, formats and media (whether known now or created in the future), to i) publish, reproduce, distribute, display and store the Contribution, ii) translate the Contribution into other languages, create adaptations, reprints, include within collections and create summaries, extracts and/or, abstracts of the Contribution, iii) create any other derivative work(s) based on the Contribution, iv) to exploit all subsidiary rights in the Contribution, v) the inclusion of electronic links from the Contribution to third party material where-ever it may be located; and, vi) license any third party to do any or all of the above.

## Disclaimer

The view(s) expressed herein are those of the author(s) and do not reflect the official policy or position of Uniformed Services University of the Health Sciences, Henry M. Jackson Foundation for the Advancement of Military Medicine, Inc., National Institutes of Health or the Department of Health and Human Services, the Defense Health Agency, the Departments of the Air Force, Navy, Army, the Department of Defense, or the U.S. Government.

Material has been reviewed by the Walter Reed Army Institute of Research. There is no objection to its presentation and/or publication. Some of the authors are employees of the U.S. Government. This work was prepared as part of official duties. Title 17, U.S.C., §105 provides that copyright protection under this title is not available for any work of the U.S. Government. Title 17, U.S.C., §101 defines a U.S. Government work as a work prepared by a military Service member or employee of the U.S. Government as part of that person’s official duties.

