## Supplemental Figures for "Demographics, Epidemiology, Mortality, and Difficult-To-Treat Resistance Patterns of Bacterial Bloodstream Infections in the Global United States Military Health System from 2010-2019: A Retrospective Cohort Study"

**Supplemental Figure 1.** Supplemental data collection diagram. A bloodstream infection (BSI) episode is a positive blood culture associated with an individual on a single unique date. The diagram is a representation of an individual with repeat bacteremia. Race and gender are fixed at the initial BSI episode. Comprehensive Ambulatory Provider Encounter Record (CAPER), Standard Inpatient Data Record (SIDR), and TriCare Encounter Data Institutional – Non-Institutional (TED I/NI) are types of data files within the Military Health System Data Repository.

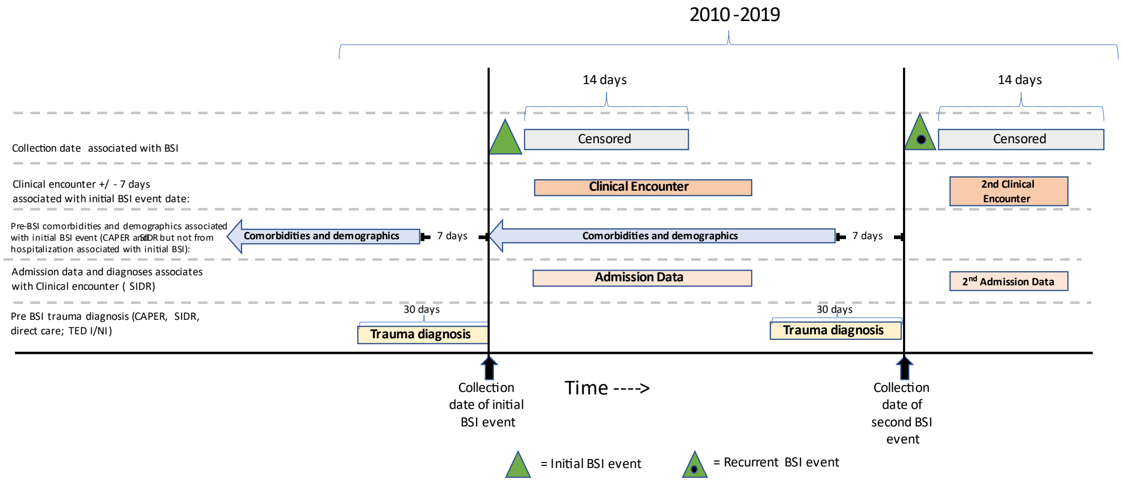

**Supplemental Figure 2. Flow diagram for Gram-positive pathogens and antimicrobial susceptibility testing.** For each species listed, multiple susceptibility testing was used so the results are not mutually exclusive.

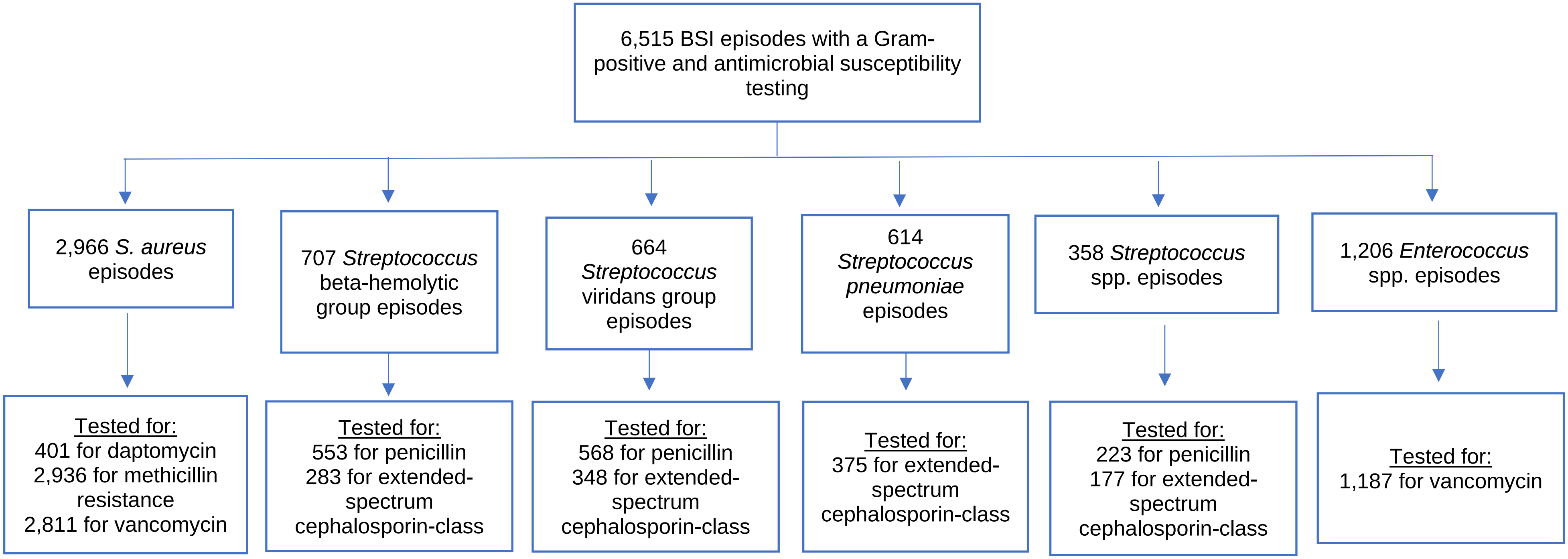
