## Supplemental Tables for "Demographics, Epidemiology, Mortality, and Difficult-To-Treat Resistance Patterns of Bacterial Bloodstream Infections in the Global United States Military Health System from 2010-2019: A Retrospective Cohort Study"

**Supplemental Table 1.** Categorization of Bacterial Subspecies that are associated with Bloodstream Infection

| **Lactose-Fermenting Gram-negative Bacilli** | ***Streptococcus* and *Enterococcus* species** | ***Staphylococcus aureus*** | **Non-Lactose-Fermenting Gram-negative Bacilli** |
|---|---|---|---|
| ***Escherichia coli*** | ***Streptococcus* β-Hemolytic group** | ***Staphylococcus aureus*** | ***Pseudomonas aeruginosa*** |
| ***Klebsiella pneumoniae*** | β-hemolytic *Streptococcus* spp. | | ***Acinetobacter* spp.** |
| ***Serratia* spp.** | β-hemolytic *Streptococcus* spp, Group A | | *Acinetobacter baumannii* |
| *Serratia ficaria* | β-hemolytic *Streptococcus* spp, Group B | | *Acinetobacter baumannii calcoaceticus* complex |
| *Serratia liquefaciens* | β-hemolytic *Streptococcus* spp, Group C | | *Acinetobacter haemolyticus* |
| *Serratia marcescens* | β-hemolytic *Streptococcus* spp, Group D | | *Acinetobacter lwoffii* |
| *Serratia odorifera 1* | β-hemolytic *Streptococcus* spp, Group F | | *Acinetobacter radioresistens* |
| *Serratia plymuthica* | β-hemolytic *Streptococcus* spp, Group G | | *Acinetobacter* spp. |
| *Serratia rubidaea* | *Streptococcus agalactiae* | | *Acinetobacter ursingii* |
| *Serratia* spp. | *Streptococcus canis* | | ***Stenotrophomonas* spp.** |
| ***Citrobacter* spp.** | *Streptococcus dysgalactiae* | | *Stenotrophomonas maltophilia* |
| *Citrobacter amalonaticus* | *Streptococcus dysgalactiae* SS. *dysgalactiae* | | |
| *Citrobacter braakii* | *Streptococcus dysgalactiae* SS. *equisimilis* | | |
| *Citrobacter farmeri* | *Streptococcus equi* | | |

| **Lactose-Fermenting Gram-negative Bacilli** | ***Streptococcus* and *Enterococcus* species** | ***Staphylococcus aureus*** | **Non-Lactose-Fermenting Gram-negative Bacilli** |
|---|---|---|---|
| *Citrobacter freundii* | *Streptococcus equisimilis* | | |
| *Citrobacter koseri (diversus)* | *Streptococcus porcinus* | | |
| *Citrobacter* spp. | *Streptococcus pyogenes* | | |
| *Citrobacter youngae* | ***Streptococcus* spp.** | | |
| ***Enterobacter* spp.** | *Streptococcus* spp. | | |
| *Enterobacter agglomerans* | *Streptococcus* nutritionally variant | | |
| *Enterobacter amnigenus* 1 | ***Streptococcus* viridans group** | | |
| *Enterobacter amnigenus* 2 | α hemolytic *Streptococcus* spp., viridans group | | |
| *Enterobacter asburiae* | *Streptococcus acidominimus* | | |
| *Enterobacter cancerogenus* | *Streptococcus alactolyticus* | | |
| *Enterobacter gergoviae* | *Streptococcus anginosus* | | |
| *Enterobacter sakazakii* | *Streptococcus constellatus* | | |
| *Enterobacter* spp. | *Streptococcus cristatus* | | |
| *Enterobacter taylorae* | *Streptococcus gordonii* | | |
| ***Proteus* spp.** | *Streptococcus mitis* | | |
| *Proteus penneri* | *Streptococcus mutans* | | |
| *Proteus* spp. | *Streptococcus parasanguinis* | | |
| *Proteus vulgaris* | *Streptococcus salivarius* | | |
| | *Streptococcus sanguinis* | | |
| | *Streptococcus sanguis* | | |
| | *Streptococcus sanguis I* | | |
| | *Streptococcus sanguis II* | | |
| | *Streptococcus sobrinus* | | |
| | *Streptococcus thermophilus* | | |
| | *Streptococcus thoraltensis* | | |

| **Lactose-Fermenting Gram-negative Bacilli** | ***Streptococcus* and *Enterococcus* species** | ***Staphylococcus aureus*** | **Non-Lactose-Fermenting Gram-negative Bacilli** |
|---|---|---|---|
| | *Streptococcus vestibularis* | | |
| | ***Streptococcus pneumoniae*** | | |
| | ***Enterococcus* species** | | |
| | *Enterococcus avium* | | |
| | *Enterococcus casseliflavus* | | |
| | *Enterococcus durans* | | |
| | *Enterococcus faecalis* | | |
| | *Enterococcus* faecium | | |
| | *Enterococcus gallinarum* | | |
| | *Enterococcus hirae* | | |
| | *Enterococcus raffinosus* | | |
| | *Enterococcus* spp. | | |

**Supplemental Table 2.** Geographic Location of Facilities Where Bloodstream Infections (BSI) Were Diagnosed

| **Geographic Location** | **BSI Episodes No. (%) (N=15,357)** |
|---|---|
| **Continental United States** | 14,653 (95.4) |
| Texas | 3128 (20.4) |
| California | 2171 (14.1) |
| Maryland | 1621 (10.6) |
| Virginia | 1272 (8.3) |
| Georgia | 971 (6.3) |
| North Carolina | 840 (5.5) |
| Florida | 532 (3.5) |
| Nevada | 403 (2.6) |
| Ohio | 327 (2.1) |
| Mississippi | 274 (1.8) |
| Kentucky | 218 (1.4) |
| District of Columbia | 202 (1.3) |
| Missouri | 202 (1.3) |
| Colorado | 160 (1.0) |
| Illinois | 71 (0.5) |
| Oklahoma | 64 (0.4) |
| Kansas | 53 (0.4) |
| Louisiana | 36 (0.2) |
| South Carolina | 34 (0.2) |
| New York | 12 (0.1) |
| Idaho | 7 (0.1) |
| Nebraska | 4 (0.03) |
| Tennessee | 2 (0.01) |
| New Mexico | 1 (0.01) |
| South Dakota | 1 (0.01) |
| **Outside of Continental United States** | |
| Hawaii | 794 (5.2) |
| Alaska | 196 (1.3) |
| Guam | 287 (1.9) |
| Germany | 194 (1.3) |
| Japan | 124 (0.8) |
| South Korea | 44 (0.3) |
| United Kingdom | 24 (0.2) |
| Italy | 15 (0.1) |
| Spain | 11 (0.1) |

| | |
|---|---|
| Cuba | 3 (0.02) |
| Bahrain | 1 (0.01) |
| Turkey | 1 (0.01) |

**Supplemental Table 3.** Comorbidities Prior to Development of Bloodstream Infections (BSI)

| **Comorbidity** | **BSI Episodes No. (%) (N=15,357)** |
|---|---|
| No comorbidity | 2,501 (16.3) |
| Chronic pulmonary disease | 7,617 (49.6) |
| Diabetes without chronic complication | 6,939 (45.2) |
| Peripheral vascular disease | 5,354 (34.9) |
| Renal disease | 5,260 (34.3) |
| Malignancy, including lymphoma and leukemia[a] | 5,238 (34.1) |
| Congestive heart failure | 4,761 (31.0) |
| Cerebrovascular disease | 4,531 (29.5) |
| Diabetes with chronic complications | 4,104 (26.7) |
| Mild liver disease | 3,391 (22.1) |
| Myocardial infarction | 2,903 (18.9) |
| Metastatic solid tumor | 1,765 (11.5) |
| Peptic ulcer disease | 1,612 (10.5) |
| Rheumatic disease | 1,372 (8.9) |
| Dementia | 1,370 (8.9) |
| Hemiplegia or paraplegia | 1,230 (8.0) |
| Moderate or severe liver disease | 653 (4.3) |
| AIDS/HIV | 89 (0.6) |
| Pre-Bloodstream Infection Updated CCI, Median score (+/- SD) | 3.0 (3.37) |

CCI – Charlson Comorbidity Index; SD – standard deviation

[a] Excludes non-melanoma malignant neoplasms of the skin

**Supplemental Table 4.** Demographics and Comorbidities Stratified by Bloodstream Infection (BSI) Organisms

| Characteristics | Total BSI Episodes, No. (%) (N=15,357) | BSI Episodes, No. (%) | | | |
|---|---|---|---|---|---|
| | | Lactose-Fermenting GNB (N=6,401) | *Streptococcus* / *Enterococcus* spp. (N=5,072) | *Staphylococcus aureus* (N=3,040) | Non-Lactose-Fermenting GNB (N=844) |
| *Age Group, years* | | | | | |
| 18-25 | 1,302 (8.5) | 431 (6.7) | 484 (9.5) | 286 (9.4) | 101 (12.0) |
| 26-35 | 1,121 (7.3) | 436 (6.8) | 388 (7.7) | 225 (7.4) | 72 (8.5) |
| 36-45 | 951 (6.2) | 391 (6.1) | 303 (6.0) | 211 (6.9) | 46 (5.5) |
| 46-64 | 4,288 (27.9) | 1,820 (28.4) | 1,381 (27.2) | 886 (29.1) | 201 (23.8) |
| ≥65 | 7,695 (50.1) | 3,323 (51.9) | 2,516 (49.6) | 1,432 (47.1) | 424 (50.2) |
| *Sex* | | | | | |
| Female | 6,811 (44.4) | 3,408 (53.2) | 2,039 (40.2) | 1,076 (35.4) | 288 (34.1) |
| Male | 8,546 (55.6) | 2,993 (46.8) | 3,033 (59.8) | 1,964 (64.6) | 556 (65.9) |
| *Race* | | | | | |
| African American | 2,524 (16.4) | 1,051 (16.4) | 859 (16.9) | 439 (14.4) | 175 (20.7) |
| Asian or Pacific Islander | 1,162 (7.6) | 608 (9.5) | 312 (6.2) | 186 (6.1) | 56 (6.6) |
| Caucasian | 8,798 (57.3) | 3,436 (53.7) | 3,022 (59.6) | 1,884 (62.0) | 456 (54.0) |
| Other | 2,873 (18.7) | 1,306 (20.4) | 879 (17.3) | 531 (17.5) | 157 (18.6) |
| *Comorbidities preceding BSI*[a] | | | | | |
| Chronic pulmonary disease | 7,617 (49.6) | 2,960 (46.2) | 2,657 (52.4) | 1,568 (51.6) | 432 (51.2) |
| Diabetes without chronic complication | 6,939 (45.2) | 2,902 (45.3) | 2,222 (43.8) | 1,464 (48.2) | 351 (41.6) |
| Peripheral vascular disease | 5,354 (34.9) | 2,023 (31.6) | 1,833 (36.1) | 1,184 (38.9) | 314 (37.2) |
| Renal disease | 5,260 (34.3) | 2,003 (31.3) | 1,762 (34.7) | 1,169 (38.5) | 326 (38.6) |

| | | | | | |
|---|---|---|---|---|---|
| Malignancy, including lymphoma and leukemia[b] | 5,238 (34.1) | 2,152 (33.6) | 1,762 (34.7) | 948 (31.2) | 376 (44.5) |
| Congestive heart failure | 4,761 (31.0) | 1,659 (25.9) | 1,712 (33.8) | 1,096 (36.1) | 294 (34.8) |
| Pre-Bloodstream Infection Updated CCI, Median score (+/- SD) | 3.0 (3.4) | 3.0 (3.3) | 3.0 (3.4) | 4.0 (3.4) | 4 (3.6) |

CCI – Charlson Comorbidity Index; GNB – Gram-negative bacilli; SD – standard deviation

[a] Most frequent comorbidities are listed. Categories are not mutually exclusive. These comorbidities are an amalgamation of the Clinical Classification Software diagnosis mapping categories.

[b] Excludes malignant neoplasms of the skin

**Supplemental Table 5.** Rates of *In Vitro* Antimicrobial Resistance of Pathogens associated with BSI Episodes

| Antimicrobial Resistance | **BSI episodes with Gram-Negative (GN) Bacteria, No. (%)** | | | | | |
|---|---|---|---|---|---|---|
| | **All GN Bacteria BSI Episodes (N=3,707)** | ***Acinetobacter* spp. (N=81)** | ***Citrobacter* spp. (N=71)** | ***Escherichia coli* (N=2,382)** | ***Klebsiella pneumoniae* (N=769)** | ***Pseudomonas aeruginosa* (N=404)** |
| Carbapenem-class antibiotic resistance | 113 (3.0) | 24 (29.6) | 3 (4.2) | 6 (0.3) | 10 (1.3) | 70 (17.3) |
| Extended-spectrum Cephalosporin-class antibiotic resistance | 400 (10.8) | 30 (37.0) | 2 (2.8) | 266 (11.2) | 63 (8.2) | 39 (9.7) |
| Fluoroquinolone-class antibiotics resistance | 865 (23.3) | 27 (33.3) | 5 (7.0) | 677 (28.4) | 79 (10.3) | 77 (19.1) |
| Piperacillin-tazobactam resistance[a] | 218 (7.9% of 2,758) | 15 (33.3% of 45) | 1 (1.8% of 56) | 85 (5.0% of 1,694) | 56 (9.6% of 583) | 61 (16.1% of 380) |
| Difficult to Treat resistance | 42 (1.1) | 19 (23.5) | 0 | 1 (0.04%) | 8 (1.0%) | 14 (3.5) |
| | **BSI Episodes with Gram-Positive Bacteria, No. (%)** | | | | | |
| | ***Staphylococcus aureus* (N=2,936)** | ***Streptococcus* β-hemolytic group (N=553)** | ***Streptococcus* spp. (N=223)** | ***Streptococcus pneumoniae* (N=375)** | ***Streptococcus* viridans group (N=568)** | ***Enterococcus* spp. (N=1,187)** |
| Methicillin resistance | 1,057 (36.0) | 0 | 0 | 0 | 0 | 0 |
| Penicillin resistance | 0 | 2 (0.4) | 55 (24.7) | 0 | 199 (35.0) | 0 |
| Vancomycin resistance | 4 (0.1% of 2,811)[b] | 0 | 0 | 0 | 0 | 205 (17.3) |
| Daptomycin resistance | 2 (0.5% of 401)[b] | 0 | 0 | 0 | 0 | 0 |
| Extended-spectrum cephalosporin class resistance | 0 | 2 (0.7 % of 283)[c] | 6 (3.4% of 177)[c] | 8 (2.1) | 20 (5.7% of 348)[c] | 0 |

[a] 949 Gram-negative isolates were not tested for piperacillin-tazobactam resistance. The proportion for each organism is calculated from the total tested.

[b] 125 *S. aureus* isolates were not tested for vancomycin resistance and 2,535 isolates were not tested for daptomycin resistance. The proportions are calculated from the total tested.

[c] 536 *Streptococcus* isolates were not tested for extended-spectrum cephalosporin class resistance. The proportions are calculated from the total tested.

**Supplemental Table 6.** Demographics and Comorbidities Among Patients Who Died Within One Year of Bloodstream Infection (BSI) Diagnosis, Stratified by BSI Organisms

| **Characteristics** | **Total Patients Who Died, No. (%) (N=2,977)** | **BSI Organism Group, No. (%)** | | | | |
|---|---|---|---|---|---|---|
| | | **Lactose-Fermenting GNB (N=922)** | ***Streptococcus / Enterococcus* spp. (N=933)** | ***Staphylococcus aureus* (N=707)** | **Non-Lactose-Fermenting GNB (N=180)** | **Multiple Organism Groups (N=235)** |
| *Age Group, years* | | | | | | |
| 18-25 | 38 (1.3) | 8 (0.9) | 15 (1.6) | 6 (0.8) | 3 (1.7) | 6 (2.5) |
| 26-35 | 52 (1.7) | 18 (1.9) | 15 (1.6) | 10 (1.4) | 4 (2.2) | 5 (2.1) |
| 36-45 | 78 (2.6) | 29 (3.1) | 18 (1.9) | 18 (2.5) | 7 (3.9) | 6 (2.5) |
| 46-64 | 607 (20.4) | 190 (20.6) | 201 (21.5) | 124 (17.5) | 37 (20.6) | 55 (23.4) |
| ≥65 | 2,202 (74.0) | 677 (73.4) | 684 (73.3) | 549 (77.6) | 129 (71.7) | 163 (69.4) |
| *Sex* | | | | | | |
| Female | 1,112 (37.3) | 383 (41.5) | 344 (36.9) | 241 (34.1) | 70 (38.9) | 74 (31.5) |
| Male | 1,865 (62.7) | 539 (58.5) | 589 (63.1) | 466 (65.9) | 110 (61.1) | 161 (68.5) |
| *Race* | | | | | | |
| African American | 499 (16.8) | 162 (17.6) | 155 (16.6) | 100 (14.1) | 37 (20.6) | 45 (19.1) |
| Asian or Pacific Islander | 213 (7.1) | 73 (7.9) | 61 (6.5) | 39 (5.5) | 21 (11.7) | 19 (8.1) |
| Caucasian | 1,869 (62.6) | 554 (60.1) | 594 (63.7) | 472 (66.8) | 106 (58.9) | 143 (60.9) |
| Other | 396 (13.3) | 133 (14.4) | 123 (13.2) | 96 (13.6) | 16 (8.9) | 28 (11.9) |
| Had ≥1 comorbidity preceding BSI | 2,889 (97.0) | 898 (97.4) | 900 (96.5) | 691 (97.7) | 175 (97.2) | 225 (95.7) |
| *Comorbidities preceding BSI*[a] | | | | | | |
| Chronic pulmonary disease | 1,801 (60.5) | 506 (54.9) | 590 (63.2) | 460 (65.1) | 103 (57.2) | 142 (60.4) |
| Malignancy, including lymphoma and leukemia[b] | 1,659 (55.7) | 554 (60.1) | 503 (53.9) | 337 (47.7) | 124 (68.9) | 141 (60.0) |
| Diabetes without chronic complication | 1,597 (53.6) | 480 (52.1) | 486 (52.1) | 426 (60.3) | 90 (50.0) | 115 (48.9) |

| | | | | | | |
|---|---|---|---|---|---|---|
| Renal disease | 1,505 (50.6) | 429 (46.5) | 489 (52.4) | 397 (56.2) | 82 (45.6) | 108 (46.0) |
| Peripheral vascular disease | 1,499 (50.4) | 423 (45.9) | 479 (51.3) | 406 (57.4) | 87 (48.3) | 104 (44.3) |
| Congestive heart failure | 1,484 (49.8) | 413 (44.8) | 470 (50.4) | 401 (56.7) | 88 (48.9) | 112 (47.7) |

GNB – Gram-negative bacilli

[a] Most frequent comorbidities are listed. Categories are not mutually exclusive. These comorbidities are an amalgamation of the Clinical Classification Software diagnosis mapping categories.

[b] Excludes malignant neoplasms of the skin
