## Supplemental Material for "Demographics, Epidemiology, Mortality, and Difficult-To-Treat Resistance Patterns of Bacterial Bloodstream Infections in the Global United States Military Health System from 2010-2019: A Retrospective Cohort Study"

### SUPPLEMENTAL MATERIAL
#### Supplemental Methods

##### Definition for Clinical Encounter
A corresponding clinical encounter for each laboratory record collection date is searched for in inpatient and outpatient military treatment facility data sources. For outpatients, a match is obtained when the collection date is within +/- 7 days of an ambulatory encounter. For inpatients, a match is obtained if the collection date occurred between the admission and discharge or if the collection date occurred within 7 days before a hospital admission. If a collection date is found in either inpatient or ambulatory data sources, a match has been found.

##### Aggregation of Microbiological Data within a BSI Episode
All Clinical and Laboratory Standards Institute (CLSI) minimum inhibitory concentrations (MIC) break points and interpretive criteria for a single BSI episode (initial or recurrent) were aggregated from any blood cultures, with growth of the same bacterial organism, from an individual on the collection date and during the preceding 13 days. If any MIC break points or interpretive criteria were determined to be either a resistant (R) or intermediate (I) phenotype, these were used to represent the aggregate antibiotic-resistant phenotype of the BSI episode.

##### Microbiological Methods Utilized at Military Treatment Facilities Laboratories
Clinical microbiology laboratories in the Military Health System predominantly use automated microbiology systems like the Vitek 2 (bioMérieux, Durham, NC), the BD Phoenix (Becton, Dickinson and Co., Franklin Lakes, NJ) or the MicroScan Walk Away (Siemens, Deerfield, NC) to identify bacteria and perform antimicrobial susceptibility testing. If certain antibiotics were not available for automated testing or per request of the provider, manual susceptibility testing including Etest and disk diffusion was performed following CLSI procedures.[1,2] Additional testing (e.g., test for the presence of carbapenemases and polymerase chain reaction [PCR] to identify the presence of resistance and virulence markers) for further characterization of the pathogens were available at a subset of laboratories.

##### Difficult-to-Treat Resistance (DTR)
The definition posed by Kadri et al.[3] was used to classify Gram-negative bacilli (GNB) as DTR. The definition required that the GNB be resistant to at least one extended-spectrum cephalosporin, one fluoroquinolone, and one carbapenem. Resistance to piperacillin-tazobactam and ampicillin-sulbactam (for *Acinetobacter baumannii* only) and resistance to aztreonam (not applicable for *A. baumannii*) were only included in the assessment of DTR when results were reported.

##### Statistical Methods for Assessing Trends in Mortality
To determine if the proportion of mortality per BSI episode changed over the study years, we fit quasi-poisson regressions by regressing the number of those who died on a time covariate, which is the calendar year minus the start year 2010 (e.g. “9” for “2019”). To account for overdispersion (e.g., data are more variable than the Poisson model assumes), log(total number of BSI episode) was used as the offset. To evaluate if there is a difference in the proportion of mortality per BSI episode between the selected antibiotic-sensitive and antibiotic-resistant bacterial species, quasi-poisson regression models were built to first examine if there is a significant trend of the mortality rate over the years among antibiotic-resistant group. If not, Fisher’s exact tests were performed by pooling data across all 10 years. Specifically, we assessed the association between overall mortality (Alive vs. Died) and

antibiotic susceptibility (susceptible vs. resistant) across 10 years. Additionally, we explored the association between overall mortality and the category of difficult-to-treat-resistant (DTR) versus non-difficult-to-treat-resistant (Non-DTR) bacteria by performing Fisher's exact tests.


#### Supplemental Results

**Bloodstream Infections Preceded by Trauma**

Trauma diagnosis preceded 1,786 (11.6%) of 15,357 BSIs with open extremity wounds (N=406, 22.7% of 1,786) most common, then open head, neck, and trunk wounds (N=395, 22.1%), and crushing or internal injury (N=312, 17.5%). Eighty (4.5%) post-trauma BSIs had burn injuries. Among the trauma-associated BSIs, *Streptococcus*/*Enterococcus* spp. were recovered from 583 (32.6%), *Staphylococcus aureus* from 514 (28.8%), lactose-fermenting Gran-negative bacilli (GNB) from 502 (28.1%), and non-lactose-fermenting GNB from 187 (10.5%) BSIs. For all organism categories, open wounds (either extremity or head, neck and/or trunk wounds) were common injury patterns preceding BSIs, ranging from 41.7% for non-lactose-fermenting GNB BSI to 15.2% for *S. aureus*. The frequency of organism categories following burn injury ranged from 2.4% for *Streptococcus/Enterococcus* spp. to 14.4% for non-lactose-fermenting GNB BSIs.

***Streptococcus* spp. Antimicrobial Resistance**

*Streptococcus* spp. BSIs (excluding β-hemolytic *Streptococcus* spp.) resistant to extended-spectrum cephalosporins ranged from 8 (2.1%) of 375 *S. pneumoniae* BSIs to 20 (5.7%) of 348 viridans *Streptococcus* BSIs.
